# Stakeholder perspectives on implementing a maternal health blended learning course for health care providers in Kenya, Nigeria and Tanzania

**DOI:** 10.64898/2026.07.29.26359229

**Authors:** Alice Norah Ladur, Uzochukwu Egere, Christopher Murray, Martin Eyinda, Onesmus Maina Muchemi, Zainab Suleiman, Mwajuma Mdoe, Maria Angelica Rweyemamu, Adacha Boslam Bello, Hauwa Mohammed, Charles Anawo Ameh

**Affiliations:** Emergency Obstetric and Quality of Care Unit, Department of International Public Health Liverpool School of Tropical Medicine. Pembrooke Place L3 5QA Liverpool United Kingdom; Liverpool School of Tropical Medicine. P.O. Box 24672 – 00100 Nairobi Kenya; The State University of Zanzibar. P.O.Box 146 Zanzibar Tanzania; Department of Clinical Nursing, University of Dodoma. P.O. Box 353 Dodoma Tanzania; Department of Obstetrics and Gyneacology, University of Dodoma. P.O. Box 353 Dodoma Tanzania; Zankli Medical Centre, No 1 Ibrahim Tahir Street, Lane 900108, Abuja, Nigeria; Department of Obstetrics and Gynaecology, University of Nairobi. P.O. Box 30197 GPO Nairobi Kenya

**Keywords:** Stakeholder involvement, stakeholder engagement, blended learning, antenatal-postnatal care, Low and Middle-Income Countries

## Abstract

**Background:** Good quality maternity care is critical in reducing maternal morbidity and mortality in regions with a high maternal and perinatal mortality. Building capacity of maternity care providers through in service training has been proven as effective in bridging the knowledge, competency, and skills gap during provision of maternity care. Meaningful involvement of stakeholder perspectives in the design and implementation of public health training programs heightens the prospects of achieving long term changes in practice and policy.

**Methods:** To explore stakeholder perspectives and experiences on the co-development of the antenatal-postnatal care course in Kenya, Nigeria, Tanzania. Data was collected through nine key informant interviews and observation notes between July – October 2022. Qualitative data was analysed in NVIVO software using inductive thematic analysis.

**Results:** Study findings showed that stakeholders were receptive of the blended learning approach for training in antenatal and postnatal care describing it as accessible, and effective in improving knowledge and skills. Course facilitators and healthcare providers valued the interactive format, opportunities for pre-session preparation, and continuous support, which contributed to improved learning experiences and collaboration.

**Conclusion:** This study highlighted the importance of involving stakeholders in the design and implementation of the antenatal-postnatal blended learning course in three countries. Eliciting feedback based on stakeholder experiences and incorporating it in real time strengthened the implementation of the blended learning course.

## Introduction

Despite a 40% reduction in the maternal mortality ratio between 2000 and 2023, sub-Saharan Africa still accounted for approximately 70% of global maternal deaths in 2023 (1). Good quality maternity care is critical in reducing maternal morbidity and mortality in sub–Saharan Africa. Improving health facility births and quality of care provided have been associated with maternal and perinatal mortality reduction (2) In their systematic review on why women die when they reach the hospital, Knight et al 2013, reported inadequate health worker training and skills mix underpinned delays leading to adverse outcomes for pregnant women and their newborns at health care facilities (3). The Lancet series on midwifery reported that 83% of maternal deaths, stillbirths and newborn health outcomes could be improved through the provision of midwifery care with the requisite skills and competencies (4). To accelerate the attainment of maternal and child-health related SDG targets, capacity strengthening of health care professionals must be effective and of the highest standards to equip them with the appropriate skills, knowledge and competencies required to provide good quality maternity care.

Healthcare professionals need regular updates in knowledge and practice of skills to remain competent. Building capacity of maternity care providers through in service training has been proven as effective in bridging the knowledge, competency, and skills gap during provision of maternity care (5). However, in-service training in Low- and Middle-Income Countries (LMICs) is predominantly delivered through face-to-face approaches, which are costly, disrupt health service delivery, and are often conducted on an ad hoc basis (6). A blended learning (BL) approach to training health care professionals is increasingly adopted in many countries to reduce prohibitive costs and disruption to service delivery in the light of severe human resource shortages in low resource settings (7). Blended learning refers to an educational approach which combines online learning and face-to-face instruction (8). Experiences from the COVID-19 pandemic underscored the importance of exploring alternatives to conventional face-to-face training methods in medical education. (9, 10). Blended learning offers an innovative approach for delivering in-service training to healthcare professionals involved in the provision of maternity care services. A randomized study of a hybrid structured skills course for clinical officers in Tanzania found that participants (n=31, 91%) believed the training would improve their ability to deliver patient-centred care, and considered the course materials, self-assessment exercises, and management case studies to be useful (11). There is evidence that a blended learning approach for in service training is as effective as face-to-face approaches in increasing providers’ knowledge (12). A quasi-experimental study found that knowledge scores were similar for the blended and conventional learning groups before training (58.5% vs 61.5%, *p* = 0.358) and three months post-training (74.7% vs 75.5% = 0.720) in basic emergency obstetric and newborn care(13). The Liverpool School of Tropical Medicine developed and piloted the antenatal-postnatal care blended learning training course for maternity care providers in Kenya, Nigeria, and Tanzania as an alternative to the previously used face-to-face training approach. (10). This study describes the perspectives of key stakeholders in the co-development and delivery of the antenatal-postnatal care blended learning course.

A stakeholder refers to an individual or group who is responsible for or affected by health- and healthcare-related decisions (14). We use the term stakeholder involvement to refer to activities undertaken by either stakeholders or researchers. In this paper, *roles* refer to stakeholders’ participation in the blended learning course design, delivery, and research activities. Stakeholders span a wide range of roles across course design, implementation, and research activities. This paper explores stakeholder perspectives of course facilitators on implementing a maternal health blended learning course in Kenya, Nigeria and Tanzania. Understanding stakeholder perspectives in in-service training helps tailor professional development to workforce needs, essential in improving quality of maternity care in low- and middle-income countries. Meaningful involvement of stakeholder perspectives in the design and implementation of public health training programs heightens the prospects of achieving long term changes in practice and or policy (15, 16).

## Materials and methods

### Study design, participants and setting

A qualitative approach nested within user experience research framework was used to explore the stakeholder perspectives and experiences involved in the development and implementation of the ANC-PNC blended learning course in Kenya, Tanzania, and Nigeria. User experience research is the systematic investigation of users’ interactions with a product, system, or service (18). It seeks to understand user behaviours, preferences, and perceptions through structured feedback gathered using both qualitative and quantitative research methods (19). It is helpful in eliciting deeper insights into user needs, perspectives, experiences, and offers opportunities to co-construct innovative design solutions with their experiences in mind (18).

The term “users” in this study includes stakeholders involved in the design and delivery of the ANC-PNC blended learning course such as project implementing partners, reproductive health coordinators, head of health facility, healthcare professionals, and course facilitators/master trainers. The stakeholder roles evolved around organization and coordination of trainings for project partners, management/key decision makers at local governance/health facility level for RH coordinators/facility heads, and course facilitators involved in midwifery training, mentorship, and provision of maternity care services respectively.

### Co-development process and procedure

This study adopted a user experience design framework proposed by Farrell 2017. The design phases followed an iterative process of discover, explore, test, and listen (19). During the discovery phase, efforts were geared towards course planning and preparation involving reviewing the literature on blended learning approaches in sexual reproductive maternal and newborn health (SRMNH) in low- and middle-income countries (LMICs). The systematic review provided valuable insights into training approaches used for in-service capacity strengthening which predominantly relied on a face-to-face approach with fewer studies adopting a blended learning format (17). The systematic review was a useful resource during the initial design phase in understanding the practical issues of implementing blended learning trainings in low resource settings. A multidisciplinary design team was constituted made up of individuals with extensive experience in midwifery practice and education including senior obstetrician and gynaecologists, midwife, nurse, paediatrician, public health specialist to spearhead the design of the blended learning (BL) course. This team subsequently identified and engaged with various stakeholders on the development of the ANC-PNC BL course. Recruitment of participants/stakeholders took place between 22/07/2022 to 04/08/2022. Data generated during this phase was gathered through recorded meetings/meeting notes and observations of the processes by one of the authors (AL).

The explore phase involved the repackaging of a four-day face-to-face training course into a three-part blended learning format; self-directed learning, facilitated virtual discussions and a two-day face-to-face session. The flipped classroom model of blended learning was used with online sessions preceding the face-to-face sessions (20). The course development phase involved reviewing literature on blended learning as a teaching approach in health care, organising planning meetings with various stakeholders and pre-testing the online platform. Planning meetings were held with various stakeholders who are subject experts in midwifery, blended learning, health education, country implementing partners and virtual learning environment (VLE) developers. The stakeholder meetings helped to ascertain content appropriate for online and face-to-face sessions. A programme schedule and course outline were drafted and shared with in-country partners for feedback and ascertain appropriateness of the course in the region. Involving in-country partners was key to ensure a local ‘buy in and sustainability of the training approach beyond the lifespan of the donor funded programme. The course content was adapted to align with ANC-PNC national guidelines and clinical practice in the three countries. The design team drew on lessons learnt from the planning meetings and literature to inform the choice of learning technologies, mode of communication and platform (e.g., Zoom to host the facilitated virtual sessions, WhatsApp for communications and the World Continuing Education Alliance (WCEA) platform to host the self-directed learning course component). The WCEA platform has provisions for offline study and can be accessed using mobile phones and computers. Courses on the WCEA platform were accepted for continuous professional development points by the health professional regulatory bodies in most sub-Saharan Africa countries. Two members of the design team peer observed LSTM colleagues delivering an online training on advanced obstetric surgical course (21). This was an invaluable development experience as it informed some of the decisions made regarding the facilitated virtual sessions on Zoom, for instance, developing of guidelines for facilitators, trainees, and orientation session for faculty to enable them to adapt to Zoom as a virtual teaching platform.

Throughout the design of the blended learning course, the design team reflected upon the intended outcomes for the online learning and face-to-face sessions and found Bloom’s taxonomy (1956) a useful resource in ensuring the course structure and content was fit for purpose (22). For instance, the design team ensured the online resources uploaded onto the VLE hosted by WCEA platform facilitated active learning, dyslexic learning needs and acquisition of knowledge with activities such as quality pre-recorded lectures, quizzes, reflective exercises, PowerPoint presentations, links for further reading and pre-post-test assessments. The Zoom sessions used case scenarios which facilitated small group discussions and opportunities to ask questions. Complex and skill-based topics were pre-allocated to the face-to-face sessions to enable health workers grasp the concepts through discussions and practice of skills. A pre-test of the self-directed learning course was carried out involving country implementing partners testing WCEA platform, course content, structure and completion time which yielded valuable feedback used to make changes prior to roll out. Data collection during this phase included observations and diary notes taken during various stakeholder meetings and pre-testing phase.

The testing phase involved the roll out of the ANC-PNC blended learning course to 89 health care providers between August – October 2022. A mixed methods design was used to assess the feasibility, change in healthcare providers’ knowledge and costs of implementing the blended learning course in Nigeria, Tanzania, and Kenya. A detailed methodology for the mixed methods study is presented elsewhere (12). Findings from this study indicated improvements in knowledge, skills and highlighted that the BL approach to ANC-PNC in-service training was feasible, cost saving compared to the face-to-face approach and acceptable to health care professionals in LMICs (12). Details about the implementer perspectives are included in this qualitative study.

In the listening phase, feedback and learning experiences were collated from all the design phases, making course adaptations, and observing impact of course adaptations/changes to learners and faculty experiences. Data was collected through key informant interviews, observations, and diary notes during debrief meetings between August – October 2022. A semi-structured topic guide was used to guide discussions in the key informant interviews which were audio recorded with permission and transcribed verbatim into English. Example questions from the key informant interviews included:

- What did you like or dislike about the blended learning training?
- What challenges did you encounter while facilitating the blended learning course?
- How will the ANC-PNC blended learning course impact healthcare practice and policy
- How should the blended learning course be delivered in future to ensure maximum impact?

An observation template was used to document experiences and processes with questions on “what worked well in each of the three blended learning components, what needed to be improved and what were the experiences on facilitating sessions”. KIIs and semi-structured observations were helpful in eliciting information on stakeholder perceptions and experiences of implementing the blended learning approach in the study sites (12). Data saturation was considered at two stages. During data collection, audio recordings and observation notes were reviewed until no new ideas or concepts emerged. During data analysis, an iterative thematic analysis was conducted, involving the review, refinement, and renaming of codes and themes until thematic saturation was achieved and a final narrative account of the findings was produced (23).

## Ethics statement

This study was conducted according to the guidelines laid down in the Declaration of Helsinki and all procedures involving research study participants were approved by Research Ethics Committees at, Liverpool School of Tropical Medicine (ID:21-052), Nigeria (Ministry of Health Oyo State: AD13/479/44511), Kenya (NACOSTI/P/21/13853), Tanzania (University of Dodoma: MA.84/261/02/‘A’/25 and Zanzibar Health Research Institute: ZAHREC/04/PR/JUNE/2022/19). A written informed consent was obtained from each participant prior to participation in the study.

### Data analysis

Qualitative data were analysed using thematic analysis in NVivo as described by Braun and Clarke (2006). Data familiarisation involved transcribing audio recordings and re-reading transcripts. Initial inductive coding was completed on all transcripts by the first author (ANL). ANL developed the coding frame/tree and updated it following discussions with co-authors. Theme development involved sorting codes into potential themes and collating all relevant coded extracts within each identified theme as part of an iterative analysis process. Themes were subsequently reviewed and refined at two levels. First, the themes were reviewed to ensure that they represented both consistent and differing views among participants/regions. Second, the themes were explored in relation to the research question to ensure that they accurately reflected the dataset. Themes and subthemes were then renamed to develop a clear and coherent narrative aligned with the study aim. The final stage of analysis involved writing a concise, and logical account of the findings across themes. Participant extracts are presented to support the key findings (24). Pseudonyms were used to maintain confidentiality in the study. Trustworthiness was achieved by applying a framework for thematic analysis; returning to the data multiple times to check for accuracy in interpretation; discussions with the study team with expertise in midwifery, blended learning, and qualitative research (25).

### Reflexivity statement

Eight authors (ANL, UE, CH, ME, OMM, ABB, HM, and CAA) work for Liverpool School of Tropical Medicine. MM and MAR are affiliated with University of Dodoma, and ZW is affiliated with State University of Zanzibar. All authors have extensive experience in qualitative research and maternity care services across Kenya, Nigeria, and Tanzania. The team comprised of six female and five male researchers. Throughout the study, the authors remained mindful of the potential influence of their professional backgrounds on the research process and took steps to minimise bias in the interview process and data interpretation. This was achieved through ongoing discussions within the research team and continuous engagement with the relevant literature. Additional information regarding the ethical, cultural, and scientific considerations specific to inclusivity in global research is included in the Supporting Information (S1 Checklist).

## Results

Table 1 describes the socio-demographic characteristics of participants in the key informant interviews. All participants were involved in the management of health facilities and provision of maternity care services in Kenya, Nigeria and Tanzania.

**Table 1.**

| Socio-demographic characteristics of participants in the key informant interviews (n=9) |  |
| --- | --- |
| Health worker cadre |  |
| Nurse | 1 |
| Nurse-midwife | 5 |
| Doctor | 3 |
| Sex |  |
| Female | 7 |
| Male | 2 |
| Clinical experience |  |
| 1-5 years | 4 |
| 6-10 years | 3 |
| 11+years | 2 |

The main findings from the study are presented under two broad themes, reflections on the process of designing the antenatal-postnatal care (ANC-PNC) blended learning course and reflections on the antenatal-postnatal care course delivery. The sub-themes include crucial preparations for the roll out of the blended learning (BL) course, acceptability of BL training approach, training time, language of instruction and ensuring change beyond training.

### Reflections on the process of designing the antenatal-postnatal care blended learning course

#### Crucial preparations prior to roll out of BL course

Preparations for training is vital for a successful implementation as well as facilitating quality educational outcomes. Prior to the launch of the blended learning course, master trainers/course facilitators were taken through a series of preparatory sessions including virtual facilitation skills, Zoom features, course materials and practice sessions.

> *“I used to wonder if we could carry out some group discussions on Zoom. Those preparation sessions on Zoom brought that to reality, and it was a good experience”* P.1.

> *“I know in our system; the Zoom meetings were new. The IT issues of clicking maybe when you want to speak, people didn’t know how to operate. Also, for the first day only, there was a bit of an issue because we were not used to being switched from one station to another. But it was rectified, and from there, things became smooth in terms of being switched from one station to the other”* P.3.

Course facilitators reported that delivering training via Zoom was a new experience, particularly the process of transitioning between breakout rooms. Nevertheless, they adapted well and successfully delivered the training. Facilitators were rotated across four breakout rooms (online stations) at twenty-minute intervals during the virtual part of the blended learning course.

#### Acceptability of blended learning training approach

The blended learning training approach was acceptable by all stakeholders in Kenya, Tanzania and Nigeria. Stakeholders loved the phased teaching approach and integration of various teaching methods for in-service training such as use of short videos, guided discussions, and practical demonstrations which was noted as important in facilitating acquisition of knowledge and skills whilst breaking the monotony of didactic lectures.

> *“What I liked most is that the participants had prior knowledge. I think when they did their self-directed learning, they were able to get some knowledge. And I think us* [facilitators] *now getting into the Zoom meeting and the face-to-face, it really made our work [teaching] a lot easier”* P.2.

> *“I appreciate this course, it helped me learn more & update my knowledge. All of them were understandable because if we got stuck in self-directed learning, we had a WhatsApp group where you get time to ask questions and you are supported, in the Zoom session part we were getting time to ask questions and receive answers, also in the face-to-face session”* P.4.

### Reflections on the ANC-PNC course delivery

#### Training time and day

Whilst the SDL part of the BL course was based on individual timing and convenience, group discussions held via Zoom required all participants to attend at scheduled afternoon times on Wednesday to Friday. We soon observed that the timing of 2-4pm was not suitable for the participants as it coincided with busy times at the health facilities and attendance fluctuated during this period, however it improved after 4pm.

> *“I observed the number of participants… to be sincere sometimes maybe you can think that somebody is there, yet maybe they are doing some other jobs. Even that one, some participants have confessed about it”* P.4.

> *“Involve the counties* [Kenya] *to really give people time for them to be able to concentrate on the Zoom meeting. What happens is sometimes somebody’s changing shift, they’re going home”* P.2.

Attendance during Zoom sessions were also affected by national health events coinciding with the training especially in Tanzania and Nigeria. Kenya had a good attendance for the three days overall however attendance was slightly poor on a Friday afternoon for personal related reasons. Zoom attendance improved in subsequent roll out of the BL course once the timing was moved to the evening hours (after 4pm) and consultations with respective country Ministries of Health for any planned activities and scheduling training clear of those days.

#### Language of instruction

The BL course was implemented in Kenya and Tanzania in East Africa and Nigeria in West Africa. Whilst the English language is widely spoken in the three countries, regional variations became noticeable during the Zoom sessions. For instance, in Nigeria and Kenya, the health workers were comfortable participating in discussions in English, whilst in Tanzania, facilitators observed that participants were struggling to communicate in English on the first day of the training. Fortunately, the facilitators/master trainers were bilingual, fluent in both English and Kiswahili and quickly switched to Kiswahili, language, participants were comfortable with and nationally spoken in Tanzania.

> *“I know in Tanzania their basic language is Kiswahili. But for us we were able to adjust very fast because we can speak both English and Kiswahili. So, when I noticed that mostly when you ask a question then they [participants] return in Kiswahili, it was a clue that as a facilitator I had to communicate in Kiswahili”* P.3.

The Zoom experiences on day one in Tanzania facilitated a transition to adapting ANC-PNC BL course into Kiswahili. The impact of this specific educational resource will reach all countries in East Africa where Swahili is considered a local language.

#### Ensuring change beyond training

To bring about lasting impact beyond short competency-based capacity strengthening initiatives, there is need to consider whole systems approach where appropriate human and financial resources as well as equipment are available in health facilities. Master trainers highlighted lack of equipment in health facilities as a challenge hindering application of new skills after training. They acknowledged the complex processes involved in the procurement of equipment/medical supplies within these regions.

> *“The challenge I heard from the health workers about PPIUD is the availability of the equipment. Because you know it requires a special forceps, a sims speculum which is not easily available in facilities*… *We know with our system, acquiring equipment may not be as easy as that”* P.6.

To address equipment challenges, master trainers facilitated brainstorming of local solutions from health workers where they pointed out inter-health facility collaboration, networking, sharing of equipment. The BL training was viewed as a collaboration platform beyond the training which could be harnessed to improve quality maternity care in the country, a win-win situation for all.

> *“…we can exchange ideas and information. I just saw some neighbouring facilities catching up. When people say-like they are lacking some equipment, then, you’ll hear somebody say, “In our facility we have so many. Come we lend you.” So, that networking also is very important because some facilities are advantaged, others are not. But through this training, we can be able to collaborate at facility level and help each other because we’re all aiming at quality care to the mother and family”* P.1.

An accessible support group was created for course facilitators and healthcare workers on WhatsApp which transitioned into peer support, collaborations between health facilities and mentorship platforms beyond the training.

## Discussion

Overall, the study found that stakeholders were receptive of the blended learning approach for training in antenatal and postnatal care describing it as accessible, and effective in improving knowledge and skills. Course facilitators and healthcare providers valued the interactive format, opportunities for pre-session preparation, and continuous support, which contributed to improved learning experiences and collaboration. The blended learning approach for in-service training has been considered favourable to health systems because it contributes to continuous professional development (CPD) and minimises disruptions to health service provision given the shortened face-to-face training component (12).

Studies have reported on the effectiveness of blended learning in enhancing knowledge acquisition among health professionals (26, 27). A similar multi country capacity building programme assessed online, virtual and in-person educational strategies highlighting significant improvements in knowledge and clinical skills in 13,000 learners in sub–Saharan Africa (28). Notable in this study, the magnitude of gains in technical knowledge and clinical confidence were observed in the face-to-face trainings compared to the virtual trainings (28). Online/blended trainings were commended for being cost effective and helpful in addressing inequities in access to training opportunities in sub–Saharan Africa (28). A qualitative study in Kenya observed that various stakeholders valued short in-service training programmes geared towards knowledge and skills acquisition and positive attitude changes for health care professionals involved in maternity care (29).

### Implications for clinical practice and policy

The findings indicate that blended learning can strengthen clinical practice by improving healthcare workers’ knowledge and ability to apply updated skills in patient care. The interactive and continuous nature of the approach supports better understanding and retention, while also encouraging collaboration, peer learning, and mentorship among clinicians. This can contribute to more consistent, evidence-based care and improved quality of services over time. For policy, the findings support integrating blended learning into national clinical training and continuing professional development frameworks. Policymakers should prioritise investment in digital training infrastructure, support the development of country-appropriate learning materials, and promote systems that enable ongoing learning and collaboration among healthcare workers. Embedding blended learning within health systems can help ensure more sustainable capacity building and long-term improvements in clinical care delivery.

### Strengths

We used an evidence-based approach (user experience research design) to co-develop and implement the ANC-PNC blended learning training programme in Kenya, Nigeria and Tanzania. A robust methodological approach embedded in the user experience framework ensured appropriate data collection and synthesis of stakeholder’s views and experiences throughout the phases of course development, implementation and evaluation. The stakeholders were invaluable in sharing technical and in-country health systems expertise which led to the effective development and implementation of the ANC-PNC blended learning course in three countries (4). The benefits of appropriate stakeholder engagement from design to evaluation of the training programme is recognised as multi-faceted including improving acceptability and feasibility of intervention, positive impacts on health outcomes, changes in policy or practice and sustainability purposes (6, 7). According to Wolcott and McLaughlin 2024, by understanding user experiences and preferences, educators can develop educational materials, resources, and activities that are engaging, motivational and region specific, ultimately improving teaching and learning outcomes (30).

### Limitations

The drawback with involving various stakeholders in the co-development of a country-specific training programme is that it requires adequate time invested in carrying out consultations, synthesising feedback, incorporating it into each phase of the course development process and evaluation as described in the methods section of the paper. However, it is worthwhile considering the long-term goal of developing acceptable, relevant and sustainable training programs like the antenatal-postnatal care blended learning training which proved to be more cost effective compared to the alternative face-to-face standalone trainings (4). Additionally, introducing something new for which health workers in low resource settings are not used to, e.g. self-directed learning and virtual facilitated learning (e.g. Zoom) could be challenging. This is something stakeholders need to be aware of whilst designing and or implementing in service capacity strengthening initiatives using the blended learning approach (31). It is worth noting that experiences reported in this study were from a small sample which may not be generalizable although they provide valuable information for persons interested in the blended learning approach for in-service training (32).

## Conclusion

This study highlighted the importance of broad stakeholder engagement in the design and implementation of the antenatal-postnatal blended learning course in three countries. Eliciting feedback based on user experiences and incorporating it in real time strengthened the implementation of the blended learning training course.

## Data Availability

All relevant data are within the manuscript and its Supporting Information files.

## Acknowledgments

We express our heartfelt gratitude to Ministry of Health, Kenya (Dr Issak Bashir); Ministry of Health, Tanzania (Dr Makuwani Ahmad Mohamed), Kaduna State Primary Health Care Development Agency (Dr Nafisat Musa Isah),; LSTM-Kenya office (Roselynne Njeri Githinji); University of Dodoma (Dr Leonard Katalambula, Mr Lalashe Kiretuni); The State University of Zanzibar (Dr Salma Abdi Mahmoud, Mr Mansab Mansab, Ms Shuwena Hamad) for the invaluable contribution in the development of the ANC-PNC blended learning course and for sharing user experiences which strengthened the course implementation and evaluation.

